# Deep learning grading of *Schistosoma japonicum*-associated liver fibrosis from ultrasound images: a weighted soft-voting ensemble of convolutional and transformer networks

**DOI:** 10.64898/2026.11.02.26364999

**Authors:** Zhaoyu Guo, Eloise S. Ockenden, Mengyan Wei, Qiang Li, Shizhen Li, Jingbo Xue, Yifeng Li, Dandan Lin, Xiaonong Zhou, Goylette F. Chami, Shizhu Li

**Affiliations:** Chinese Center for Disease Control and Prevention (Chinese Center for Tropical Diseases Research), National Key Laboratory of Intelligent Tracking and Forecasting for Infectious Diseases, National Institute of Parasitic Diseases, National Health Commission Key Laboratory of Parasite and Vector Biology, WHO Collaborating Centre for Tropical Diseases, National Center for International Research on Tropical Diseases, Shanghai 200025, China; Big Data Institute, Nuffield Department of Population Health, University of Oxford, Oxford OX3 7LF, United Kingdom; School of Global Health, Chinese Center for Tropical Diseases Research, Shanghai Jiao Tong University School of Medicine, Shanghai 200025, China; Jiangxi Province Key Laboratory of Schistosomiasis Prevention and Control, Nanchang 330096, China

## Abstract

**Background:** Despite progress towards schistosomiasis elimination in China, *Schistosoma japonicum*-associated liver fibrosis remains a source of morbidity. Ultrasound grading is subjective, and experienced sonographers are scarce in endemic counties.

**Methods:** We developed a deep learning ensemble to grade fibrosis from a single B-mode ultrasound image without manual region-of-interest delineation. We analysed 2,921 images from 2,921 adults (aged 18–88 years; median age 54; 46.4% female) at a schistosomiasis hospital in Jiangxi Province, China (2019–2023). Grades 0, 1, 2 and 3 accounted for 12.9%, 34.6%, 32.4% and 20.2% of images, respectively. Reference grades were derived from clinical ultrasound records and radiologist reassessment using the Chinese Handbook of Schistosomiasis Control, with disagreements resolved by consensus. Predictions from ResNet-18, EfficientNet-B0, DenseNet-121 and Swin-T were combined by validation-weighted soft voting. Performance was evaluated on a held-out test set of 440 images.

**Results:** The ensemble achieved a macro-F1 of 0.8527 (95% CI 0.8184–0.8856), accuracy of 0.8500 (95% CI 0.8136–0.8818) and quadratic weighted kappa of 0.9187 (95% CI 0.8983–0.9373) for four-grade classification, outperforming each individual network in macro-F1. All ensemble misclassifications involved adjacent grades. For detecting significant fibrosis (grades 2–3; test-set prevalence 53.0%), sensitivity was 0.9700, specificity 0.9179, positive predictive value 0.9300, negative predictive value 0.9645 and AUROC 0.9817.

**Conclusions:** This single-image approach may assist automated grading of *S. japonicum*-associated liver fibrosis in endemic settings. External validation is needed before clinical implementation.

## 1 Introduction

Schistosomiasis-related morbidity persists in China despite substantial progress towards transmission interruption and the marked decline in active infections. Schistosomiasis affects more than 250 million people worldwide, while *Schittotoma japonicum* is endemic mainly in China and the Philippines, with small residual foci in Indonesia. Compared with other major human schistosomes, *Schittotoma japonicum* has substantially greater fecundity, resulting in a high tissue egg burden and potentially severe hepatosplenic morbidity [1]. Chronic infection can progress to hepatic fibrosis and portal hypertension and, in severe cases, to advanced schistosomiasis with massive splenomegaly, ascites, colonic proliferative lesions, severe growth retardation or gastroesophageal variceal bleeding [2]. As China moves towards schistosomiasis elimination and active infections become increasingly rare, residual morbidity is becoming concentrated among previously infected populations[3–5]. Importantly, hepatic fibrosis may continue to progress in some individuals even after antiparasitic treatment and interruption of transmission, although the mechanisms underlying this post-treatment progression remain incompletely understood [6]. By the end of 2025, 26,688 patients with advanced schistosomiasis remained registered across China’s 450 historically endemic counties, even though 421 of these counties had achieved elimination and the remaining 29 had reached transmission interruption [7]. In Changshu, Jiangsu Province, a follow-up study found that 75 previously infected individuals still developed advanced schistosomiasis, although transmission had been interrupted for more than 35 years [6].

Accurate grading and longitudinal monitoring of liver fibrosis in these individuals are therefore important needs of current control programmes [8]. Two-dimensional grey-scale (B-mode) ultrasonography is non-invasive, repeatable and widely available, and can visualise the characteristic hepatic changes of schistosomiasis japonica. A distinctive feature is interseptal fibrosis, in which fibrous septa between hepatic lobules appear as echogenic bands outlining polygonal areas and producing a characteristic fish-scale or network pattern. Under the Chinese ultra-sound grading scheme used in endemic areas, hepatic fibrosis is classified from grade 0 to 3. Grade 0 represents a normal liver; grade 1 is characterised by scattered focal echodense areas and mildly coarsened echoes; grade 2 by stronger echogenic bands forming a fish-scale or fine network pattern; and grade 3 by a coarse, contiguous echogenic network, often accompanied by more extensive parenchymal and portal abnormalities [9]. These grades represent a continuum of increasing echogenicity, coarsening and network formation rather than categories separated by a single discrete feature. Visual grading is therefore inherently observer-dependent, particularly at the boundaries between adjacent grades [9, 10].

The grades used in this study follow the national handbook criteria in place when the images were read [11]. In 2025 the National Institute of Parasitic Diseases convened more than 30 Chinese and international experts to publish an expert consensus on the ultrasound diagnostic pathway for this condition [9]. That consensus explicitly identifies artificial intelligence-assisted image recognition, grading and follow-up monitoring as an important direction for the field. It is aligned with the WHO(World Health Organization) framework for verifying interruption of transmission and for post-interruption surveillance [9].

Artificial intelligence applied to ultrasound imaging has begun to show promise for automated assessment of schistosomal liver fibrosis. Guo et al. extracted 851 radiomic features from manually delineated regions of interest and trained support vector machines, achieving validation AUCs of 0.771–0.834 for discrimination between adjacent grades [11]; this approach requires manual delineation before feature extraction and formulates grading as six pairwise tasks rather than a single four-grade prediction. Ockenden et al. developed SchistoTrackNet to classify Niamey liver-image patterns of *Schittotoma mantoni*-associated periportal fibrosis in Uganda [12]; the disease phenotype and label space differ from those of *S. japonicum*, and no validated mapping exists between the Niamey patterns and the Chinese four-grade scale [13]. Most directly relevant, Xu et al. trained a single ResNet-50 on 167,702 images from 9,588 participants at 20 centres to predict a 36-level ordinal refinement of the four clinical grades [26]. That study establishes that deep learning can grade *S. japonicum*-associated fibrosis at scale [14].

We therefore asked whether the four national grades could be predicted from a single B-mode image, without manual region-of-interest delineation, using images from one county hospital in Jiangxi Province, China (2,921 patients, one image each, 2019–2023). Because the number of images was small by the standards of deep learning, we combined four backbones from different architectural families (ResNet-18, EfficientNet-B0, DenseNet-121 and Swin-T) by weighted soft voting rather than enlarging a single network, with weights determined exclusively from validation-set performance before evaluation on the held-out test set. We also asked where the gain of the ensemble comes from, and report an activation map for each backbone so that the image evidence behind a prediction is available to the reading clinician.

## 2 Methods

### 2.1 Study population

This was a retrospective analysis of abdominal ultrasound images acquired at the Second People’s Hospital of Duchang County, Jiangxi Province, China, between 2019 and 2023. Jiangxi Province is a major schistosomiasis-endemic area of China, and residents of the Poyang Lake region are at risk of exposure to infested water. The hospital is the designated local hospital for schistosomiasis diagnosis and treatment. Schistosomiasis was diagnosed according to the national Diagnostic Criteria for Schistosomiasis (WS 261–2006)[15], and *Schittotoma japonicum*-associated liver fibrosis was graded with reference to the Handbook of Schistosomiasis Control[16].

The inclusion criteria were evidence of current or previous *Schittotoma japonicum* infection, defined as a documented history of schistosomiasis, a positive serological test for anti-*Schittotoma japonicum* antibodies, or a positive parasitological stool examination for *S. japonicum*, together with availability of right subcostal and right subcostal oblique ultrasound images. Cases with conditions that could affect ultrasound assessment, such as fatty liver, hepatitis, or drug- or alcohol-induced liver injury, were excluded, as were images with severe distortion or poor quality.

Images were acquired with a Philips EPIQ 5 ultrasound system and a C5-1 convex-array transducer at 3.5–5.0 MHz, exported in DICOM format and de-identified before modelling. The ultrasound grade recorded in the medical record was retrieved, and a radiologist with more than 5 years of experience in schistosomiasis ultrasonography re-evaluated each image. A second radiologist reviewed cases with discordant grades, and disputed cases were discussed to decide on inclusion and final grade. The final consensus labels were used for model training and defined a four-class task: schistosomal liver fibrosis grades 0, 1, 2 and 3 (G0–G3). The grades are distinguished by the coarseness and spatial organisation of the parenchymal echo. Grade 0 shows a uniform liver parenchymal echo with fine or slightly thicker bright spots. In grade 1 the parenchymal echo is uneven and the bright spots become coarser. In grade 2 the echo is unevenly distributed with enhanced bright spots that form a fish-scale pattern, and a fine mesh-like echo is seen throughout the liver [11]. In grade 3 the echo is uneven with thicker, more intense bright spots and higher-pitched echoes, and the echoes form a densely packed, coarse network-like high-echo band [11].

### 2.2 Ethics

This study was reviewed and approved by the Ethical Review Committee of the National Institute of Parasitic Diseases, Chinese Center for Disease Control and Prevention (approval no. 2021019). All participants were informed of the study’s aims, procedures, potential benefits and risks before enrolment, and each provided written informed consent; no minors were enrolled. The present analysis was performed retrospectively on de-identified ultrasound images, with no additional intervention or contact with participants.

### 2.3 Data partitioning and pre-processing

The 2,921 included images, with one image per patient, were split into training, validation and test sets at a ratio of approximately 70:15:15, stratified by fibrosis grade using a random seed of 42. Once defined, the training, validation and test sets were kept fixed throughout model development, with the test set held out and used only for the final evaluation. The proportion of each fibrosis grade differed by no more than 0.47 percentage points across the three subsets (Fig. 1a).

**Figure 1:**
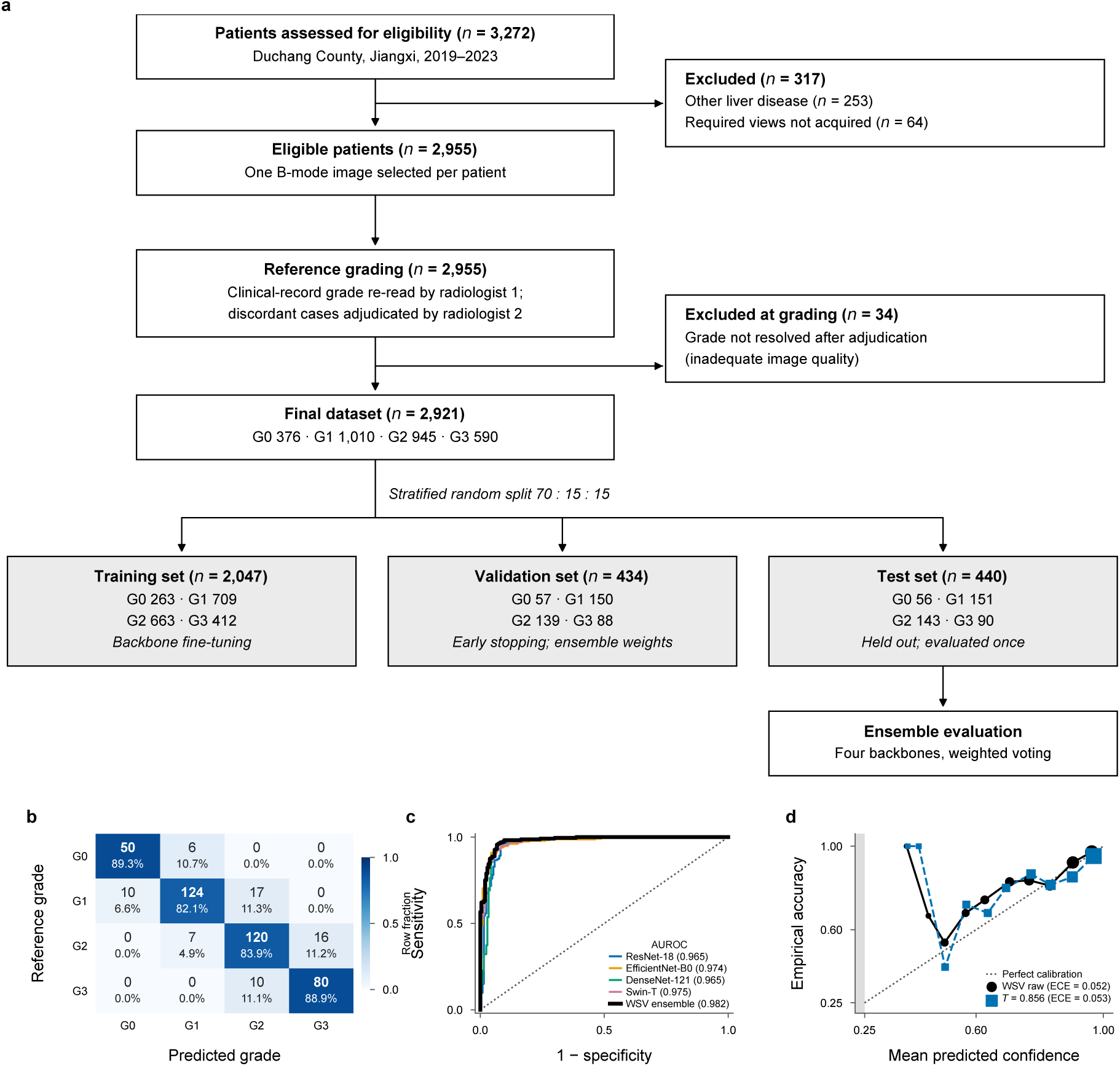
Cohort composition, error structure and calibration on the fixed test set (*n* = 440). **a,** Flow of participants. Of 3,272 patients with a documented history of *Schistosoma japonicum* infection examined at Duchang County Second People’s Hospital between 2019 and 2023, 317 were excluded before image selection and 34 at reference grading, leaving 2,921 patients with one B-mode image each. The dataset was split at the patient level, stratified by grade, into training (n = 2,047), validation (n = 434) and test (n = 440) sets. The training set was used to fine-tune the four backbones, the validation set for early stopping and for determining the ensemble weights, and the test set for a single final evaluation. Numbers in each set are given by grade (G0–G3). **b,** Confusion matrix of the weighted soft-voting (WSV) ensemble; cell values are numbers of images and the shading is the fraction of each reference-grade row. No cell lies more than one grade off the diagonal. c, Receiver operating characteristic curves for the significant-fibrosis (grade ≥2) task, scored as P(G2) + P(G3); the legend gives the area under the curve for each model. d, Reliability curve of the ensemble before and after temperature scaling (*T* = 0.856 fitted on the validation set).

Images were stored in Digital Imaging and Communications in Medicine (DICOM) format as single-channel grey-scale arrays of 360 × 385 pixels. Each image was linearly stretched to 0–255 using its own minimum and maximum grey levels and resized to 224 × 224 pixels by bilinear interpolation. It was then converted to a tensor and normalised with a mean of 0.5 and a standard deviation of 0.5. Per-image stretching was intended to make the model rely on the relative grey-level distribution rather than on absolute echogenicity and thus reduce the effect of different gain settings. During training, we applied random horizontal flipping (p = 0.5) and random affine transformation (rotation ±10^◦^, translation ±5%, scaling 0.95–1.05). No augmentation was applied during validation or testing.

### 2.4 Models and ensemble

Models were trained and evaluated on a single workstation with a 20-core Arm CPU and one NVIDIA GB10 accelerator sharing 120 GB of unified memory (Linux kernel 6.17, NVIDIA driver 580.142). The implementation used Python 3.12.3 with PyTorch 2.11.0 and torchvision 0.26.0 built against CUDA 13.0; metrics were computed with scikit-learn and activation maps with the grad-cam package. All package versions are recorded in Supplementary Information.

We chose four backbones from different architectural families: ResNet-18 (residual convolutional network), EfficientNet-B0 (compound-scaled convolutional network with channel activation), DenseNet-121 (densely connected convolutional network) and Swin-T (hierarchical vision Transformer) [17–20]. The main challenge in four-grade assessment is the discrimination of adjacent grades, which represent a continuum of ultrasonographic changes and have shown lower discriminability than more widely separated grades in previous work [11]. Different architectures encode distinct inductive biases: convolutional networks impose stronger locality and spatial-equivariance priors, whereas self-activation allows information to be integrated across more spatially distributed image regions [21]. Swin-T further combines hierarchical representations with shifted-window activation to enable information exchange across neighbouring windows [20]. These architectural differences may result in partially non-overlapping prediction errors. Probability-level averaging can therefore exploit complementary classifier outputs and reduce errors specific to individual models [22].

All four backbones were trained on identical data with an identical training procedure – loss function, optimiser, learning-rate schedule, augmentation and stopping rule – and differed only in architecture. Each was initialised from ImageNet-1K pre-trained weights; the first convolutional (or patch-embedding) layer was adapted to single-channel input by averaging its pre-trained RGB kernels, and the whole network was then fine-tuned [23]. The loss was class-weighted cross-entropy with label smoothing[24] (coefficient 0.05), the weight of each class being proportional to the inverse of its training-sample count and normalised to sum to the number of classes. We used the AdamW optimiser (Adam with decoupled weight decay)[25] with a learning rate of 1 × 10^−4^ and a weight decay of 1 × 10^−4^, linear warm-up over the first 3 epochs and cosine annealing thereafter. The batch size was 32, gradients were clipped to a global norm of 1.0, and training ran for up to 30 epochs. Training stopped early if the validation macro-F1 did not improve for 8 consecutive epochs, and the weights with the highest validation macro-F1 were retained as the final model. The random seed was fixed at 42, with cuDNN deterministic mode enabled and benchmarking disabled to improve run-to-run consistency.

Ensembling was performed at the probability level. Each backbone produced predicted class probabilities for the four grades; these were combined linearly with weights fixed before any test-set evaluation, and the grade with the highest combined probability was the final prediction: *P* (*g*) = Σ*_a_ w_a_* · *P_a_*(*g*). The weights were the normalised best validation macro-F1 of each backbone, *w_a_* = F1*_a_/*Σ*_b_* F1*_b_*: 0.2453 for ResNet-18, 0.2525 for EfficientNet-B0, 0.2480 for DenseNet-121 and 0.2542 for Swin-T. No test data were used to estimate the weights. Unlike majority voting, probability weighting preserves each model’s uncertainty across grades, which suits a four-grade task in which adjacent grades are easily confused.

### 2.5 Model performance

The primary metric was the macro-averaged F1 score (macro-F1). Secondary metrics were accuracy, quadratically weighted kappa (QWK), mean absolute error (MAE) and, for the binary task of significant fibrosis (grade ≥2), the area under the receiver operating characteristic curve (AUROC), sensitivity and specificity. On the fixed test set (n = 440), 95% confidence intervals (CIs) were estimated by non-parametric bootstrap with 1,000 resamples (random seed 42). Models were compared by paired bootstrap, in which the same resampling indices were applied to the predictions of each model and to the reference labels. Stability was assessed by five-fold cross-validation.

### 2.6 Activation map

Grad-CAM (gradient-weighted class activation mapping) was computed for all four backbones on their final spatial feature layer. For ResNet-18, EfficientNet-B0 and DenseNet-121, Grad-CAM was applied to the final convolutional feature maps. For Swin-T, the output of the second layer normalisation in the final transformer block was reshaped from 49 spatial tokens to its native 7 × 7 grid before Grad-CAM computation. For all backbones, the target class was the grade predicted by the ensemble so that the resulting maps referred to the same class for a given image. Maps were upsampled to 224 × 224 pixels, smoothed with a Gaussian kernel (σ = 1.5), and normalised to [0, 1] within each image. We then calculated the equal-weighted mean activation, the population standard deviation across the four maps, and the number of backbones with normalised activation above 0.5 at each pixel. The diversity index was defined as the image-wide mean of the standard-deviation map, and the agreement ratio as the proportion of pixels at which all four backbones exceeded 0.5.

## 3 Results

### 3.1 Study population

Of 3,272 patients examined between 2019 and 2023, 317 were excluded before image selection (other liver disease, *n* = 253; required views not acquired, *n* = 64) and 34 at reference grading, leaving 2,921 patients with one image each (Fig. 1a). The mean age was 51.7 years (SD 14.8), and 53.6% were male and 46.4% were female. Grades G0–G3 accounted for 376 (12.9%), 1,010 (34.6%), 945 (32.4%) and 590 (20.2%) patients, and the distribution differed by no more than 0.5 percentage points across the training, validation and test sets.

### 3.2 Grading performance of the individual backbones and the ensemble

Table 1 summarises the performance of the four backbones and the weighted soft-voting (WSV) ensemble on the fixed test set (*n* = 440; metrics are defined in Methods). The four backbones performed similarly, with test macro-F1 from 0.798 to 0.807 and overlapping 95% confidence intervals. ResNet-18 had the highest macro-F1 (0.807; 95% CI, 0.769–0.846), Swin-T the highest QWK (0.890; 95% CI, 0.863–0.912) and Swin-T had the highest QWK (0.890) and grade ≥ 2 fibrosis AUROC (0.975) among the individual models.

**Table 1:**
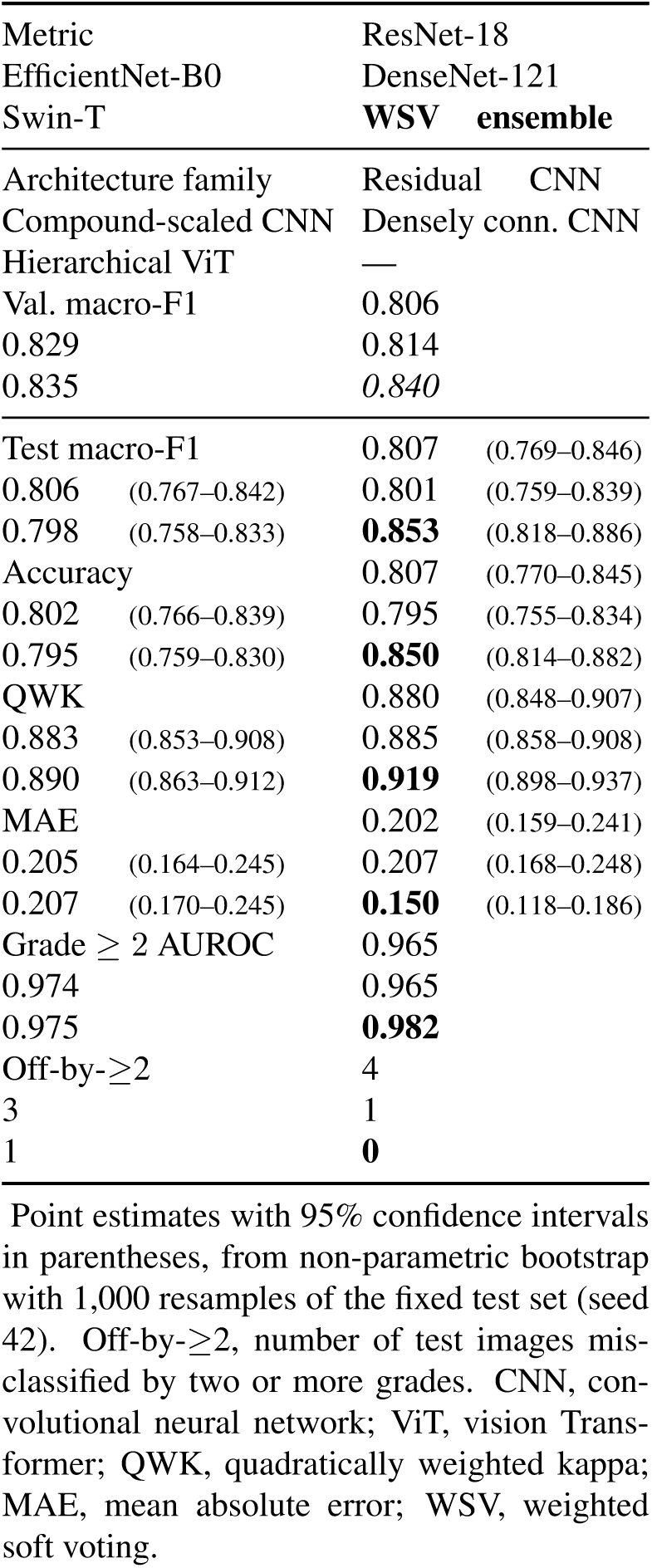
Grading performance of the individual backbones and the weighted soft-voting ensemble on the fixed test set (*n* = 440).

| Metric |  | ResNet-18 |  |
|---|---|---|---|
| EfficientNet-B0 |  | DenseNet-121 |  |
| Swin-T |  | **WSV ensemble** |  |
| Architecture family |  | Residual CNN |  |
| Compound-scaled CNN |  | Densely conn. CNN |  |
| Hierarchical ViT |  | — |  |
| Val. macro-F1 |  | 0.806 |  |
| 0.829 |  | 0.814 |  |
| 0.835 |  | *0.840* |  |
| Test macro-F1 |  | 0.807 | (0.769–0.846) |
| 0.806 | (0.767–0.842) | 0.801 | (0.759–0.839) |
| 0.798 | (0.758–0.833) | **0.853** | (0.818–0.886) |
| Accuracy |  | 0.807 | (0.770–0.845) |
| 0.802 | (0.766–0.839) | 0.795 | (0.755–0.834) |
| 0.795 | (0.759–0.830) | **0.850** | (0.814–0.882) |
| QWK |  | 0.880 | (0.848–0.907) |
| 0.883 | (0.853–0.908) | 0.885 | (0.858–0.908) |
| 0.890 | (0.863–0.912) | **0.919** | (0.898–0.937) |
| MAE |  | 0.202 | (0.159–0.241) |
| 0.205 | (0.164–0.245) | 0.207 | (0.168–0.248) |
| 0.207 | (0.170–0.245) | **0.150** | (0.118–0.186) |
| Grade ≥ 2 AUROC |  | 0.965 |  |
| 0.974 |  | 0.965 |  |
| 0.975 |  | **0.982** |  |
| Off-by-≥2 |  | 4 |  |
| 3 |  | 1 |  |
| 1 |  | **0** |  |

Point estimates with 95% confidence intervals in parentheses, from non-parametric bootstrap with 1,000 resamples of the fixed test set (seed 42). Off-by-≥2, number of test images misclassified by two or more grades. CNN, convolutional neural network; ViT, vision Transformer; QWK, quadratically weighted kappa; MAE, mean absolute error; WSV, weighted soft voting.

The WSV ensemble reached a macro-F1 of 0.853 (95% CI, 0.818–0.886), accuracy of 0.850 (0.814–0.882), QWK of 0.919 (0.898–0.937), MAE of 0.150 (0.118–0.186) and grade ≥ 2 fibrosis AUROC of 0.982, the best value on every metric. In the paired bootstrap, in which the same resampled images were scored for both models, the ensemble exceeded ResNet-18 in macro-F1 by 0.046 (95% CI, 0.019–0.073), EfficientNet-B0 by 0.047 (0.020–0.074), DenseNet-121 by 0.052 (0.021–0.084) and Swin-T by 0.055 (0.024–0.088); none of the intervals included zero. The ensemble also made smaller errors. Every one of its 66 misgraded images was assigned a grade adjacent to the reference grade, whereas the single backbones misgraded 1 to 4 images by two grades or more (Table 1).

The confusion matrix of the ensemble (Fig. 1b) shows how the errors were distributed. The proportion of images assigned their reference grade was 89.3% for G0 (50 of 56), 82.1% for G1 (124 of 151), 83.9% for G2 (120 of 143) and 88.9% for G3 (80 of 90). All six G0 errors were graded G1 and all ten G3 errors were graded G2, as the extreme grades can only be missed in one direction. For the two intermediate grades, errors went in both directions but more often upwards: of the 27 G1 errors, 17 were graded G2 and 10 were graded G0, and of the 23 G2 errors, 16 were graded G3 and 7 were graded G1. G1 was the hardest grade for the ensemble and for three of the four backbones, with per-grade sensitivity from 0.689 (Swin-T) to 0.828 (EfficientNet-B0).

Among the single models, EfficientNet-B0 had the highest F1 for G0 (0.842) and G1 (0.836), ResNet-18 for G2 (0.788) and Swin-T for G3 (0.836). The ensemble had the highest F1 in all four grades (0.862, 0.861, 0.828 and 0.860). Its gain over the individual backbones was largest for G2, where the paired difference was significant against three of the four backbones after Benjamini–Hochberg correction across the 16 grade-by-backbone comparisons; in total, 8 of the 16 comparisons were significant.

Table 2 gives the positive and negative predictive values (PPV, NPV), sensitivity and specificity of each model for each grade, computed one grade against the other three pooled. NPV exceeded 0.90 in 15 of the 20 model-by-grade combinations; the five exceptions were at G1 and G2. PPV varied more than NPV, spanning 0.699 to 0.919 against 0.855 to 0.986. Across the table, a high PPV went with a low sensitivity rather than a high one (*r* = −0.51, *P* = 0.02), the expected trade-off between assigning a grade liberally and conservatively. G1 shows this most clearly, with the highest mean PPV of any grade (0.889) and the lowest mean sensitivity (0.768). The ensemble had the best PPV for G0, G2 and G3 and the best NPV for G2 and G3. For G3 it combined a sensitivity of 0.889 with a PPV of 0.833, so advanced cases were largely detected and, when detected, usually correct. These predictive values depend on the grade mix of the test set (13% G0, 34% G1, 33% G2, 20% G3) and would differ in a population with another prevalence.

**Table 2:** Positive and negative predictive values by fibrosis grade.

| Model | G0 (*n* = 56) | G1 (*n* = 151) | G2 (*n* = 143) | G3 (*n* = 90) |
|---|---|---|---|---|
| *CNN-based* |  |  |  |  |
| *ResNet-18* |  |  |  |  |
| PPV | 0.7424 | **0.9194** | 0.7378 | 0.8256 |
| NPV | 0.9813 | 0.8829 | 0.9203 | 0.9463 |
| Sensitivity | 0.8750 | 0.7550 | **0.8462** | 0.7889 |
| Specificity | 0.9557 | **0.9654** | 0.8552 | **0.9571** |
| *EfficientNet-B0* |  |  |  |  |
| PPV | 0.8276 | 0.8446 | 0.7895 | 0.7426 |
| NPV | 0.9791 | **0.9110** | 0.8762 | 0.9558 |
| Sensitivity | 0.8571 | **0.8278** | 0.7343 | 0.8333 |
| Specificity | **0.9740** | 0.9204 | 0.9057 | 0.9257 |
| *DenseNet-121* |  |  |  |  |
| PPV | 0.8214 | 0.8692 | 0.7403 | 0.7700 |
| NPV | 0.9740 | 0.8774 | 0.8986 | 0.9618 |
| Sensitivity | 0.8214 | 0.7483 | 0.7972 | 0.8556 |
| Specificity | **0.9740** | 0.9412 | 0.8653 | 0.9343 |
| *Transformer-based* |  |  |  |  |
| *Swin-T* |  |  |  |  |
| PPV | 0.6986 | 0.9043 | 0.7582 | 0.7980 |
| NPV | **0.9864** | 0.8554 | 0.9059 | 0.9677 |
| Sensitivity | **0.9107** | 0.6887 | 0.8112 | 0.8778 |
| Specificity | 0.9427 | 0.9619 | 0.8754 | 0.9429 |
| *Ensemble* |  |  |  |  |
| *WSV ensemble* |  |  |  |  |
| PPV | **0.8333** | 0.9051 | **0.8163** | **0.8333** |
| NPV | 0.9842 | 0.9109 | **0.9215** | **0.9709** |
| Sensitivity | 0.8929 | 0.8212 | 0.8392 | **0.8889** |
| Specificity | **0.9740** | 0.9550 | **0.9091** | 0.9543 |

A summary of the positive predictive value, negative predictive value, sensitivity and specificity achieved by each model for each fibrosis grade, computed one grade against all others pooled. The highest value across the five models for each grade and each measure is emboldened.

We also read the four-grade output as two clinical yes/no questions. For *any fibrotit* (G1– G3 versus G0; 384 versus 56 images), the ensemble detected 97.4% of fibrotic images with a specificity of 89.3% and an AUROC of 0.983; the single models ranged from 94.3% to 97.4% in sensitivity and from 82.1% to 91.1% in specificity. For *tignificant fibrotit* (G2–G3 versus G0– G1; 233 versus 207 images) the ensemble achieved an AUROC of 0.982 (Fig. 1c), above every single model (0.965–0.975). At the operating point given by the predicted grade, the ensemble’s sensitivity was 97.0%, specificity 91.8%, PPV 93.0% and NPV 96.4%. Three backbones matched its sensitivity of 97.0% but with lower specificity (86.5–88.4%); EfficientNet-B0 was more specific (92.3%) but less sensitive (93.6%).

To test whether these results depended on the particular training run, we repeated the entire pipeline with 15 training seeds on the same fixed split, changing only the random seed. Ensemble macro-F1 was 0.848 ± 0.011 across seeds (range, 0.820–0.862). The single backbones varied more, with seed-to-seed SDs from 0.014 to 0.020. The ensemble exceeded the best single backbone in all 15 seeds, by 0.025 on average.

We also re-partitioned the whole pool of 2,921 images into five patient-level folds and retrained the four backbones and the ensemble in each fold (Methods). Ensemble macro-F1 on the held-out folds was 0.832 ± 0.014, against 0.811 ± 0.011 for the best single backbone of each fold. Because the folds re-partition the same pool of patients as the fixed split, this measures the stability of the pipeline rather than providing an independent validation.

### 3.3 Activation patterns of the backbones

Grad-CAM maps show where the evidence for a backbone’s decision was concentrated. We refer to them as activation maps. We first asked whether the four backbones highlight the same part of an image. For each image we took the hottest 10% of pixels in each map and measured the overlap between every pair of backbones. Two regions of that size placed independently would overlap by 0.10; identical regions would give 1. The observed medians were 0.25 to 0.51 across the six pairs. We also measured the distance between the two maps’ single hottest pixels. This was a median of 41 to 68 pixels, or 18 to 30% of the frame width, against 114 pixels for two independently placed points. The maps therefore overlap far more than chance without coinciding (Fig. 2).

**Figure 2:**
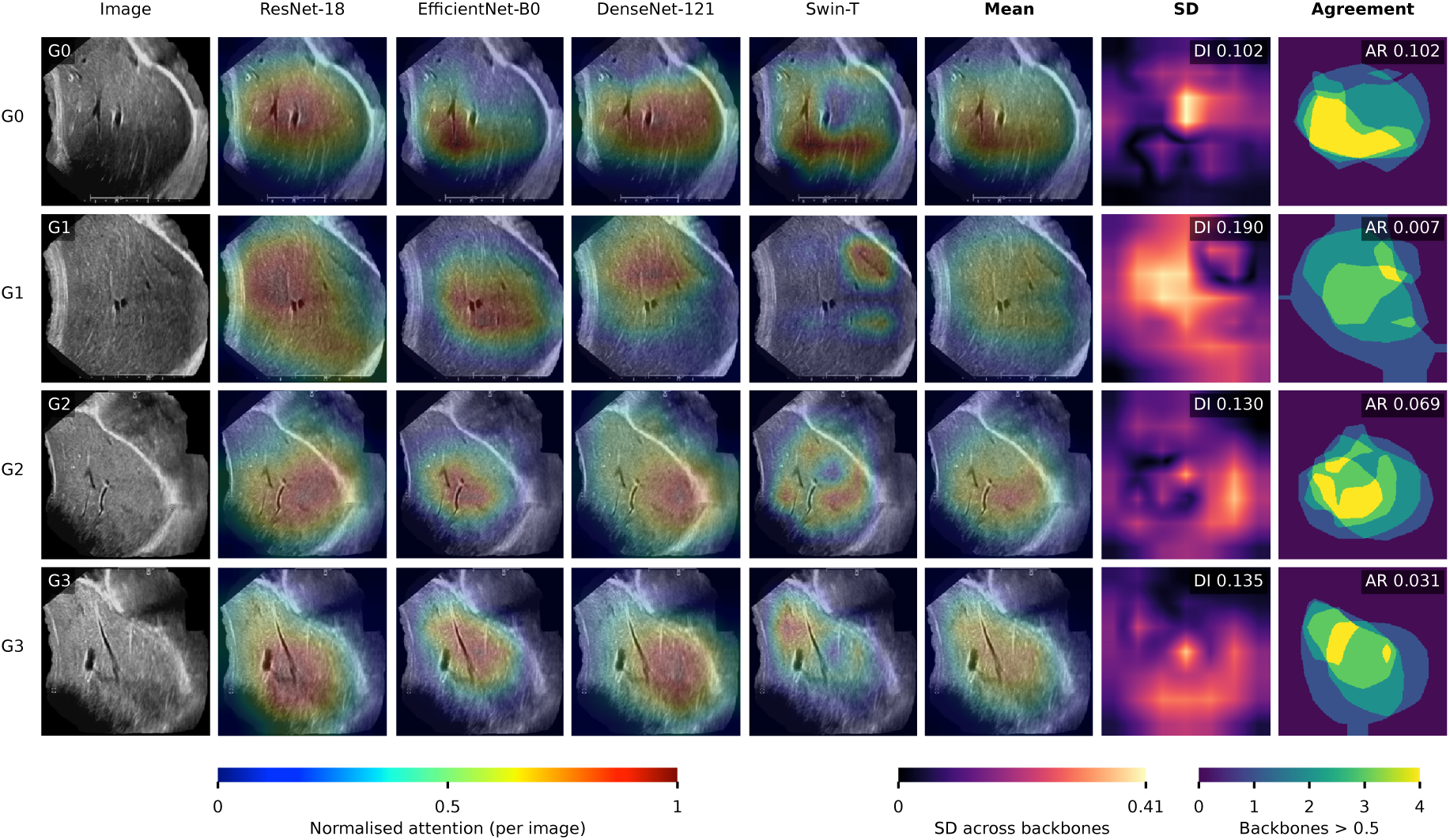
Per-pixel comparison of backbone activation. Each row shows, for one grade (G0–G3), a high-confidence correctly classified test case. Columns: input image; Grad-CAM maps of ResNet-18, EfficientNet-B0, DenseNet-121 and Swin-T, normalised per image to [0, 1]; equal-weighted mean of the four maps; standard deviation (SD) across backbones; and number of backbones with normalised activation above 0.5 (agreement). DI, diversity index (image-wide mean of the SD map); AR, agreement ratio (proportion of pixels at which all four backbones exceed 0.5).

The differences between maps are not an artefact of how the maps were produced. The target layer of every backbone outputs a 7 × 7 grid for a 224 × 224 input (Methods), and all four maps were upsampled to the input size by the same operator and normalised the same way, so no backbone’s map is coarser or smoother than another’s by construction. Nor is the ordering explained by map extent. Swin-T does mark a smaller area, with the region above half of a map’s own maximum covering 9% of the frame against 29% for ResNet-18.

The disagreement followed architecture. Ranking the six pairs by overlap, the three pairs of convolutional backbones came above all three pairs containing the transformer. The most similar pair was ResNet-18 with EfficientNet-B0, and the least similar was ResNet-18 with Swin-T. We averaged the three pairs of each kind within each image and tested the paired difference. The convolutional pairs were the more similar on all six measures we computed, three in the main text and three in the Supplementary Information, all with *P <* 1 × 10^−12^ (Wilcoxon signed-rank, *n* = 420). For Swin-T, we also evaluated attention rollout and gradient-weighted rollout (Supplementary Information).

These maps show that the four backbones focus on partly different image regions. They do not prove that the models are correct, and without lesion-level annotations we do not assign the highlighted areas to specific anatomical structures.

### 3.4 Source of the ensemble gain

The ensemble gain did not come from correcting errors shared by all four backbones. All 35 cases (8.0%) misclassified by every backbone were also misclassified by the ensemble, with a mean posterior probability of only 0.105 for the reference grade. Conversely, no case was correct for all four backbones but wrong for the ensemble. The gain arose only when the members disagreed: in 24 cases (5.5%), at least two backbones were wrong but the ensemble was correct, including 2 cases in which three backbones were wrong (Fig. 3). In case `G1_925`, the errors were spread over two grades, and the reference grade prevailed after averaging with a posterior probability of 0.394. On the whole test set, the macro-F1 of the WSV ensemble (0.853) was higher than that of majority (hard) voting (0.832). Of the 20 cases on which the two methods disagreed, soft voting was correct in 14 and wrong in 6.

**Figure 3:**
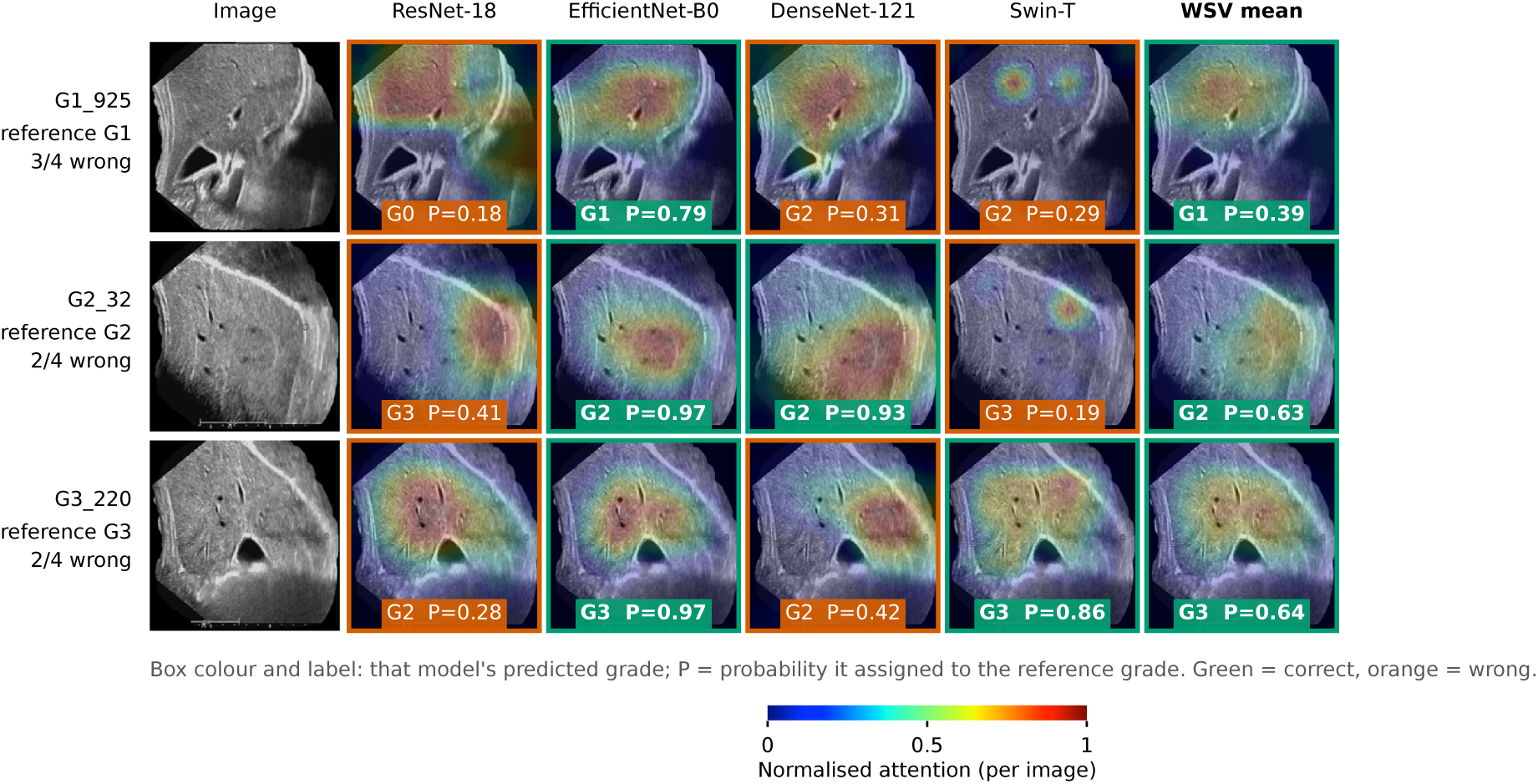
Grad-CAM activation for cases in which the ensemble corrected backbone errors. Each row shows one test case (case identifier, reference grade and number of erring backbones at left). Columns: input image; Grad-CAM map of each backbone; WSV-weighted mean map. The label in each panel gives the grade predicted by that model and the probability (P) it assigned to the reference grade. Green frames, correct predictions; orange frames, incorrect predictions.

## 4 Discussion

Grading schistosomal liver fibrosis on B-mode ultrasound is the examination on which surveillance and follow-up of previously infected people in endemic areas of China depend, and it is read by eye, with disagreement concentrated at the boundaries between adjacent grades [9, 10]. Using 2,921 images, we trained four backbones of different architectural families and combined them by weighted soft voting into a single four-grade reader that takes the whole image as input, without manual delineation of a region of interest. On the fixed test set the ensemble reached a macro-F1 of 0.853 (95% CI, 0.818–0.886), and its advantage over each backbone (0.046 to 0.055 in macro-F1) had a paired 95% confidence interval that excluded zero. The advantage held in all 15 training seeds and five cross-validation folds, so it does not depend on one fortunate training run. The ensemble also reduced the size of the errors. In the reported run, every one of its errors was between adjacent grades, whereas each backbone misgraded 1 to 4 images by two grades or more.

### Is the ensemble worth its cost, and how it could be deployed?

The ensemble improved macro-F1 significantly over the best single backbone, a gain that readers can weigh against the cost of running four networks. Three considerations favour the ensemble in this application. First, the gain is consistent: it appeared in every seed and in 8 of the 16 grade-by-backbone comparisons after correction for multiple testing, most reliably at G2. Second, the ensemble reduced the errors that matter most clinically, those spanning two or more grades, in the reported run and in most seeds. Third, and contrary to what the word ensemble suggests, the ensemble does not require a larger machine than a single network. The reason is that the four backbones are independent and are combined only through their output probabilities, so they can be evaluated one after another, each loaded, run and released before the next. Peak memory is then set by the largest member rather than by their sum. We measured this on the deployed configuration, at the input size the pipeline uses. Evaluated sequentially, the ensemble’s peak accelerator memory was 131.4 MB, identical to that of Swin-T alone, its largest member, and below the 219.1 MB measured when all four were held in memory at once. What the ensemble costs is time, not hardware: 163.3 ms per image on four CPU threads, against 80.4 ms for the slowest single backbone and 10.8 ms for the fastest, and 10.9 ms per image with all four resident on an NVIDIA GB10 accelerator. The four deployed backbones total 49.7 million parameters, 1.8 times the 27.5 million of Swin-T alone and about twice the 23.5 million of a single ResNet-50 in the same configuration. Because images are graded one at a time rather than in a real-time stream, a few hundred milliseconds is not a clinical constraint, and no additional class of hardware is needed.

Two deployment routes therefore follow from the same set of trained weights, and the choice between them is governed by data protection rather than by computing capacity. In the first, a single backbone runs locally on the ultrasound workstation, so that no image leaves the department and no network connection is required. On these data ResNet-18 had the highest macro-F1 among the single models, while EfficientNet-B0 offers the best balance of specificity and size, at 4.0 million parameters and 10.8 ms per image. The cost is the ensemble gain reported above, and the loss of the reduction in errors spanning two or more grades. In the second, the full ensemble runs on an institutional server, which keeps one auditable copy of the model, allows it to be updated and monitored centrally, and removes any dependence on the specification of the individual workstation. This route requires images to be transmitted, so it is the more demanding of the two in terms of governance: it presupposes de-identification at source, an encrypted channel, and the data-protection approvals of the institutions involved.

### The need for ensemble-based approaches

Combining classifiers at the probability level reduces the errors that are specific to individual members while leaving shared errors untouched [22], and this is what we observed. All 35 test images misgraded by every backbone were also misgraded by the ensemble; on these images the four posteriors agreed on a wrong grade and the reference grade received a mean probability of only 0.105, so averaging could not recover it. The gain came entirely from the images on which the backbones disagreed: in 24 of them, two or three backbones were wrong and the ensemble was right, either because a confident minority outweighed a less confident majority or because the errors fell on different grades and the reference grade had the highest probability after averaging. For this mechanism to work, the members must err on different images, and members of different architectural families are the natural candidates, because convolutional networks and vision transformers are known to build different internal representations of the same image [21]. The activation maps were consistent with the observed prediction diversity. Convolutional backbones showed more similar Grad-CAM patterns to one another than to Swin-T across all six similarity measures, indicating partly different spatial attribution patterns across architectures. These differences may be related to the complementary prediction errors observed in the ensemble, but Grad-CAM alone cannot establish a causal link between attribution differences and ensemble performance.

### Relation to previous work

Two lines of work are relevant. Machine-learning readers for *Schittotoma mantoni* periportal fibrosis [12, 13] address a different pattern, the echogenic thickening along the portal tracts of the Niamey protocol, whereas *S. japonicum* produces a parenchymal network pattern graded by its coarseness and organisation [8, 9]. For *S. japonicum* itself, Xu and colleagues recently reported a single ResNet-50, trained on 167,702 images from 9,588 participants at 20 centres, that predicts a 36-level ordinal refinement of the four clinical grades [14]. Theirs rests on the scale and the granularity of the labelled data, whereas ours asks how much can be obtained from 2,921 images acquired at a single centre. That question led us to combine backbones of different architectural families rather than to enlarge a single one. It also led us to report an activation map for every backbone, so that the image evidence on which a prediction rests is available to the reading clinician alongside the grade itself. Their trained model is not released, and we did not attempt a direct comparison of the two. Additionally, the study by Xu and colleagues required 18 ultrasound images per participant, whereas the present study requires only a single, relatively easy-to-acquire cross-sectional image of the liver.

### Limitations

This study has several limitations. First, it was a single-centre retrospective study with a relatively homogeneous sample, and the external generalisability of the model has not been validated. Second, only the clearest ultrasound image of each patient was included; information from multiple planes and time points was not used, and whether it could further improve grading remains to be evaluated. Third, the model used grey-scale images alone, without clinical variables such as serology and medical history or multimodal information such as elastography. Fourth, the study population was relatively old, with a mean age of 51.7 years, which may limit the generalisability of the model to younger populations. This age distribution likely reflects the changing epidemiology of schistosomiasis in endemic areas, where new infections have declined substantially and most newly infected individuals recover after treatment, reducing the repeated infections that historically contributed to progression to advanced disease.

Future work will address these limitations along three lines. A multicentre prospective validation study is ongoing to assess the stability and generalisability of the model across devices, populations and clinical settings. Where necessary, federated learning may support joint modelling without sharing raw images. We will also use multi-frame or multi-plane images as input and integrate clinical variables and elastography to enrich the information available to the model. Finally, we will extend the model to longitudinal monitoring to track the progression or regression of fibrosis in individual patients.

## Conclusion

A weighted soft-voting ensemble of four backbones graded *Schittotoma japonicum*-associated liver fibrosis from a single B-mode ultrasound image on the four-grade scale, achieving a macro-F1 of 0.853 on the original test partition. Its performance was higher than that of each individual backbone, and in the reported run all misclassifications were confined to adjacent grades. The observed improvement was associated with complementary errors across the four architecturally different backbones. For detecting grade ≥2 fibrosis, the ensemble achieved an AUROC of 0.982. These findings support further evaluation of automated fibrosis grading in endemic settings, particularly through external and prospective validation.

## Data Availability

The ultrasound images analysed in this study are derived from patient data and are not publicly available because of privacy and ethical restrictions. De-identified data are available from the corresponding author (S.L.) on reasonable request, subject to applicable institutional and ethical requirements.

## 5 Data availability

The ultrasound images analysed in this study contain patient information and are not publicly available; de-identified data are available from the corresponding author (S.L.) on reasonable request.

## 6 Code availability

The training, evaluation, duplicate-audit and figure-generation code, together with the exact split manifests, is available from the corresponding author (S.L.) on reasonable request.

## 7 Acknowledgements

We are grateful to the Second People’s Hospital of Duchang County and the National Institute of Parasitic Diseases, Chinese Center for Disease Control and Prevention, for their support of this study. We also gratefully acknowledge the support of the China Scholarship Council (CSC) through a postdoctoral fellowship.

## 8 Author contributions

Zhaoyu Guo: Conceptualization, Methodology, Software, Validation, Formal analysis, Investigation, Data curation, Visualization, Writing – original draft, Funding, Writing – review & editing. Eloise S. Ockenden: Methodology, Validation. Mengyan Wei: Investigation, Data curation. Qiang Li: Investigation, Data curation. Shizhen Li: Investigation, Data curation. Jingbo Xue: Investigation, Data curation. Xiaonong Zhou: Funding, Conceptualization, Supervision, Project administration, Writing – review & editing. Goylette F. Chami: Conceptualization, Methodology, Supervision, Project administration, Writing – review & editing. Shizhu Li: Funding, Conceptualization, Methodology, Supervision, Project administration, Writing – review & editing. All authors reviewed and approved the final manuscript.

